# Associations of socioeconomic position and adverse childhood experiences with health-related behaviour changes and changes to employment during the first COVID-19 lockdown in the UK

**DOI:** 10.1101/2021.05.18.21257397

**Authors:** Madeleine L Smith, Annie Herbert, Amanda Hughes, Kate Northstone, Laura D Howe

**Author notes:** Corresponding author: Madeleine L Smith, Oakfield House, Oakfield Grove, Clifton, Bristol, BS8 2BN, UK.

## Abstract

**Background:** Non-pharmaceutical interventions to reduce the spread of COVID-19 may have disproportionately affected already disadvantaged populations.

**Methods:** We analysed data from 2710 young adult participants of the Avon Longitudinal Study of Parents and Children. We assessed the associations of socioeconomic position (SEP) and Adverse Childhood Experiences (ACEs, e.g. abuse, neglect, measures of family dysfunction) with changes to health-related behaviours (meals, snacks, exercise, sleep, alcohol and smoking/vaping), and to financial and employment status during the first UK lockdown between March-June 2020.

**Results:** Experiencing 4 or more ACEs was associated with reporting decreased sleep quantity during lockdown (OR 1.53, 95% CI: 1.07-2.18) and increased smoking and/or vaping (OR 1.85, 95% CI: 0.99-3.43); no other associations were seen between ACEs or SEP and health-related behaviour changes. Adverse financial and employment changes were more likely for people with low SEP and for people who had experienced multiple ACEs; e.g. people who had been in the ‘never worked or long-term unemployed’ or ‘routine and manual occupation’ categories pre-lockdown were almost 3 times more likely to have stopped working during lockdown compared with people who were in a higher managerial, administrative or professional occupation pre-lockdown (OR 2.83, 95% CI: 1.45-5.50 and OR 2.68, 95% CI: 1.63-4.42 respectively).

**Conclusion:** Adverse financial and employment consequences of lockdown were more likely to be experienced by people who have already experienced socioeconomic deprivation or childhood adversity, thereby widening social inequalities. Despite this, in this sample of young adults, there was little evidence that lockdown worsened inequalities in health-related behaviours.

**BOX:** *What is already known on this subject?:*

- Non-pharmaceutical interventions implemented to mitigate the spread of COVID-19 (e.g. nationwide lockdown) have affected the lifestyles and livelihoods of many, with potential consequences for physical health and financial wellbeing.
- Existing evidence suggests sociodemographic inequalities in the effects of lockdown, but details of the factors influencing these inequalities remain unclear.

*What this study adds?:*

- A history of adverse childhood experiences may be associated with decreased sleep and increased smoking/vaping during lockdown. SEP and ACEs were not associated with changes in other health-related behaviours in this cohort of young adults.
- Adverse financial and employment consequences of lockdown were more likely to be experienced by people who have already experienced socioeconomic deprivation or childhood adversity, thereby widening social inequalities. Our findings highlight a need for continued support of people who experience ACEs into adulthood, and demonstrate that this need may have increased during the COVID-19 pandemic.

## INTRODUCTION

Measures taken to reduce the spread of COVID-19 in the United Kingdom (UK), including a national ‘lockdown’ implemented on 23^rd^ March 2020,[1] have had profound effects on everyday life. Concerns that lockdown measures could worsen existing socioeconomic and health inequalities by disproportionately affecting disadvantaged populations were expressed early on in the pandemic.[2–5] Since then, evidence suggests these concerns were well founded.[6] Determining who is more likely to have suffered adverse effects will be critical in guiding comprehensive and targeted support for those most in need.

Several studies have highlighted consequences of lockdown on physical and mental health.[7–15] During lockdown, health behaviours such as diet, alcohol consumption, smoking, sleep and physical activity were subject to rapid change, with both increases and decreases observed. Analysis of five British cohort studies suggested lockdown widened socioeconomic inequalities in sleep but did not affect inequalities in other health behaviours.[7] Financial and employment changes were also prevalent during lockdown, due in part to the effective shutdown of a number of sectors, including hospitality and travel.[16,17] Initial UK evidence suggested younger people were more affected by loss of employment and income during lockdown.[18] There was a clear social gradient for loss of income and employment, and difficulties accessing food and medicines, with individuals of lower socioeconomic position (SEP) more negatively affected.[19,20]

Impact on individuals of the UK lockdown may also differ by other factors, including a history of adverse childhood experiences (ACEs), that is, stressful events in childhood such as abuse, neglect, and family dysfunction. Exposure to ACEs has been shown to be associated with worse health and health-related behaviours across adolescence and adulthood.[21–23] Therefore, individuals with a history of ACEs may be less resilient to the damaging effects of periods of adversity such as lockdown. Whether ACEs predict increased hardship during lockdown remains to be determined.[24]

Using data from the Avon Longitudinal Study of Parents and Children (ALSPAC), a UK prospective cohort study collected during the COVID-19 pandemic,[25] we investigated associations of SEP and ACEs with health and financial consequences of the first UK lockdown (March-June 2020).

## METHODS

### Study population

The study used data from the Avon Longitudinal Study of Parents and Children (ALSPAC), an ongoing birth cohort study that recruited 14,541 pregnant women in Avon, UK with expected delivery dates between 1st April 1991 and 31st December 1992.[26–28] Mothers, children, and mother’s partners have been followed up using clinics, questionnaires, and linkage to routine data. Additional eligible cases were recruited to the study when the oldest participants were approximately 7 years old. Including these, the offspring cohort consists of 14,901 participants that were alive at one year of age. The website contains details of available data through a fully searchable data dictionary: http://www.bristol.ac.uk/alspac/researchers/our-data/. Ethical approval was obtained from the ALSPAC Law and Ethics Committee and the Local Research Ethics Committees. Participants provided written informed consent.

This study is based on 2710 offspring participants who responded between 26^th^ May and 5^th^ July 2020 to a questionnaire rapidly deployed early on during the COVID-19 pandemic.[25] The questionnaire was developed and deployed using REDCap (Research Electronic Data CAPture tools), a secure web application for online data collection hosted at the University of Bristol.[29]

### ACEs

ACE measures were derived from questions relating to multiple forms of ACEs reported by participants and their mothers at multiple timepoints from birth to 23 years of age. Full details are described elsewhere.[30] Briefly, dichotomous indicators of exposure between 0 and 16 years were created for the ten ACEs included in the World Health Organisation ACE international questionnaire.[31] The ten ACEs we considered were:

1. ever sexually abused or forced to perform sexual acts or touch someone in a sexual way (sexual abuse)
2. adult in family was ever physically cruel towards or hurt the child (physical abuse)
3. parent was ever emotionally cruel towards the child or often said hurtful/insulting things to the child (emotional abuse)
4. child always felt excluded, misunderstood, or never important to family, parents never asked or never listened when child talked about their free time (emotional neglect)
5. parent was a daily cannabis or any hard drug user or had an alcohol problem (parental substance abuse)
6. parent was ever diagnosed with schizophrenia or hospitalised for a psychiatric problem or, during the first 18 years of the child’s life, parent had an eating disorder (bulimia or anorexia), used medication for depression or anxiety, attempted suicide, or scored above previously established cut-offs for depression (Edinburgh Postnatal Depression Scale (EPDS) >12–13) (parental mental illness or suicide)
7. parents were ever affected by physically cruel behaviour by partner or ever violent towards each other, including hitting, choking, strangling, beating, and shoving (violence between parents)
8. parents separated or divorced (parental separation)
9. child was a victim of bullying on a weekly basis (bullying)
10. parent was convicted of a crime (parental criminal conviction)

Based on the sum of the dichotomous ACE constructs, each participant was given an ACE score (0-10 ACEs), which was categorised as 0, 1, 2-3, or 4+ ACEs for comparability with previous studies.

### SEP

Socioeconomic position was indicated by occupational social class at age 23 (∼4 years prior to the start of the pandemic), using the 3-class National Statistics Socio-economic classification (NS-SEC)[32]:

1. Higher managerial, administrative, and professional occupations (NS-SEC group 1)
2. Intermediate occupations (NS-SEC group 2)
3. Routine and manual occupations (NS-SEC group 3)
4. Never worked and long-term unemployed (LTU)

This was derived from participant self-reports of occupation, business/industry and job responsibilities from a questionnaire administered at mean age 23 years. Responses were coded into the first three NS-SEC categories. Participants were instructed to skip certain questions if they were “not engaged in any form of work”, so participants who skipped these questions only created the fourth category. Participants who indicated they were full-time students were excluded from analysis of SEP (Supplementary Figure 1).

### Outcomes

The term lockdown refers to the stay-at-home order made by the UK government on Monday 23^rd^ March 2020. A questionnaire deployed between 26^th^ May and 5^th^ July 2020 asked participants (mean age 27.8 years) to report whether each of the following activities had decreased a lot, decreased a little, stayed the same, increased a little or increased a lot since lockdown: number of home-cooked meals eaten, number of meals eaten in a day, number of snacks eaten in a day, amount of physical activity/exercise, amount of sleep, alcohol and smoking/vaping. We combined the responses to give three levels: “decreased”, “stayed the same” and “increased”.

Participants could select “not applicable” if they didn’t do the activity before lockdown and weren’t doing it at the time of questionnaire completion. For the first five variables (number of home-cooked meals, number of meals, number of snacks, amount of physical activity, amount of sleep), the number of participants selecting “not applicable” was negligible (n=6, 4, 8, 2, 4, respectively) so these responses were coded as missing. For alcohol and smoking, “not applicable” was coded as a fourth category, representing non-drinkers and non-smokers/vapers.

In the same questionnaire, participants reported their employment situation before lockdown. We categorised responses into working and not working pre-lockdown, and combined these with employment status at questionnaire completion to derive change in employment status during lockdown. This had five categories: employed with no change, employed with reduced hours, employed and on furlough or paid or unpaid leave, no longer working, and not working pre-lockdown.

Participants also reported how their financial situation compared to before the COVID-19 pandemic. Possible responses were: “much worse off”, “a little worse off”, “about the same”, “a little better off”, or “much better off”; we grouped the first two into ‘worse off’ and the last two into ‘better off’. Participants reported whether they or their partner had made any new claims for benefits since the pandemic, or had used rent or mortgage or other debt holidays since the pandemic.

### Missing data

ACE measures were derived from >500 questions answered between birth and 23 years of age, and no participant had data on all these questions. We therefore used multivariate multiple imputation to estimate missing values. This avoids exclusion of participants whilst reducing selection bias. Participants were only excluded from analyses if they responded to <10% of ACE questions (Supplementary Figure 1). For participants who responded to ≥50% of questions for an ACE, these questions were used to create a binary indicator of presence/absence of that ACE. For participants who responded to <50% of questions for an ACE, presence/absence of the ACE was set to missing and was imputed. The ACE score was derived after imputing presence/absence of individual ACEs. Given known sex differences in ACE prevalence, missing data for males and females were imputed separately, and the datasets re-combined before analysis. The imputation model included all outcomes, exposures and covariates included in analysis, and 24 auxiliary variables likely to predict missingness or ACE exposure (details in Supplementary File). Some participants had missing SEP data, so we included in the imputation model 8-class NS-SEC[32], from which the 4-group version described above was derived. Since only participants who completed the COVID-19 questionnaire were included in analysis, most of the analytic sample had complete outcome data. For those that had not responded to certain sections or questions, these were imputed in the same model. Using the mice package[33] in R version 4.0.2, we created 50 imputed datasets for both males and females, with 10 iterations per dataset. Imputed values were combined using Rubin’s rules[34] and trace plots used to check convergence of estimates.

### Statistical analyses

All analysis was conducted in R version 4.0.2. To explore how participants in our analysis differed from the full cohort, we compared the distribution of maternal education between included participants and those excluded due to missing data. We used multinomial logistic regression to examine associations of ACE score, individual ACEs, and SEP with health behaviour and employment outcomes during lockdown. We assessed unadjusted associations, and associations adjusted for covariates. These included participant’s ethnicity, age in years at questionnaire completion, and home ownership. We also adjusted for their mother’s marital status, parity, and age at the participant’s birth, and educational qualifications of the mother and their partner. We tested for an interaction between each exposure and sex on the outcome.

## RESULTS

### Participant characteristics

For associations of ACEs with outcomes, 2707 participants were included in analyses. For associations of SEP with outcomes, 2557 participants were included in analyses (Supplementary Figure 1). Between 0-16 years, 81.4% of participants were exposed to at least one ACE, and 20.1% to 4 or more ACEs (Table 1). Prevalence of individual ACEs ranged from 6% for sexual abuse to 43.7% for parental mental illness or suicide (Table 1). Of participants included in the SEP analyses, 44% were employed in higher managerial, administrative, and professional occupations, and 9.2% were long-term unemployed at age 23 (Supplementary Table 1). Participants included in analysis had more highly educated mothers than participants excluded due to missing data (Supplementary Table 2).

**Table 1.** Participant characteristics. Characteristics of the participants included in ACE analyses using data from multi-variate multiple imputation. N=2707.

|  |  | **n (%)** |
| --- | --- | --- |
| **ACE score** | 0 | 504 (18.6) |
|  | 1 | 679 (25.1) |
|  | 2-3 ACEs | 980 (36.2) |
|  | 4+ | 544 (20.1) |
| **physical abuse** |  | 622 (23.0) |
| **sexual abuse** |  | 163 (6.0) |
| **emotional abuse** |  | 659 (24.3) |
| **emotional neglect** |  | 529 (19.5) |
| **bullying** |  | 646 (23.9) |
| **violence between parents** |  | 604 (22.3) |
| **parental substance abuse** |  | 330 (12.2) |
| **parental mental illness or suicide** |  | 1183 (43.7) |
| **parental criminal conviction** |  | 280 (10.3) |
| **parental separation** |  | 780 (28.8) |
| **number of homecooked meals** | decreased | 104 (3.8) |
|  | stayed the same | 909 (33.6) |
|  | increased | 1694 (62.6) |
| **number of meals** | decreased | 280 (10.3) |
|  | stayed the same | 1942 (71.7) |
|  | increased | 485 (17.9) |
| **number of snacks** | decreased | 358 (13.2) |
|  | stayed the same | 1023 (37.8) |
|  | increased | 1326 (49.0) |
| **exercise quantity** | decreased | 1087 (40.2) |
|  | stayed the same | 520 (19.2) |
|  | increased | 1100 (40.6) |
| **sleep quantity** | decreased | 630 (23.3) |
|  | stayed the same | 1141 (42.1) |
|  | increased | 936 (34.6) |
| **alcohol quantity** | decreased | 462 (17.1) |
|  | stayed the same | 828 (30.6) |
|  | increased | 1047 (38.7) |
|  | non-drinker | 370 (13.7) |
| **smoking/vaping quantity** | decreased | 103 (3.8) |
|  | stayed the same | 287 (10.6) |
|  | increased | 299 (11.0) |
|  | non-smoker/vaper | 2018 (74.5) |
| **change in employment status** | employed no change | 1587 (58.6) |
|  | employed reduced hours | 261 (9.6) |
|  | furlough/paid leave/unpaid leave | 496 (18.3) |
|  | stopped working during pandemic | 165 (6.1) |
|  | not working pre-pandemic | 198 (7.3) |
| **financial situation since pandemic** | worse off | 685 (25.3) |
|  | stayed the same | 1012 (37.4) |
|  | better off | 1010 (37.3) |
| **claimed benefits since pandemic** |  | 336 (12.4) |
| **mortgage/rent holiday since pandemic** |  | 181 (6.7) |

Changes to health-related behaviours during lockdown varied in direction (Table 1; Supplementary Table 1). For example, 40.2% of participants reported a decrease in exercise quantity and 40.6% an increase, while 23.3% reported a decrease in sleep quantity and 34.6% an increase.

A large proportion of participants (58.6%) reported no change in employment during lockdown, 9.6% were working reduced hours, 18.3% were on furlough or unpaid leave and 6.1% had stopped working during lockdown (Table 1). 25.3% of participants reported feeling financially worse off compared to pre-lockdown, whereas 37.3% felt financially better off (Table 1).

There was no evidence for interactions between each exposure and sex on any of the outcomes.

### Associations of SEP and ACEs with health-related behaviours during lockdown

Participants who were in NS-SEC group 3 or LTU at age 23 had higher odds of both decreased and increased number of meals, and higher odds of decreased sleep quantity, compared with participants in NS-SEC group 1 (Table 2; Supplementary Table 3). LTU participants were less likely to have either increased or decreased their physical activity compared to NS-SEC 1 participants. However, odds ratios were imprecisely estimated, and confidence intervals spanned the null. For other health-related behaviours, there was little evidence of socioeconomic differences.

**Table 2:**
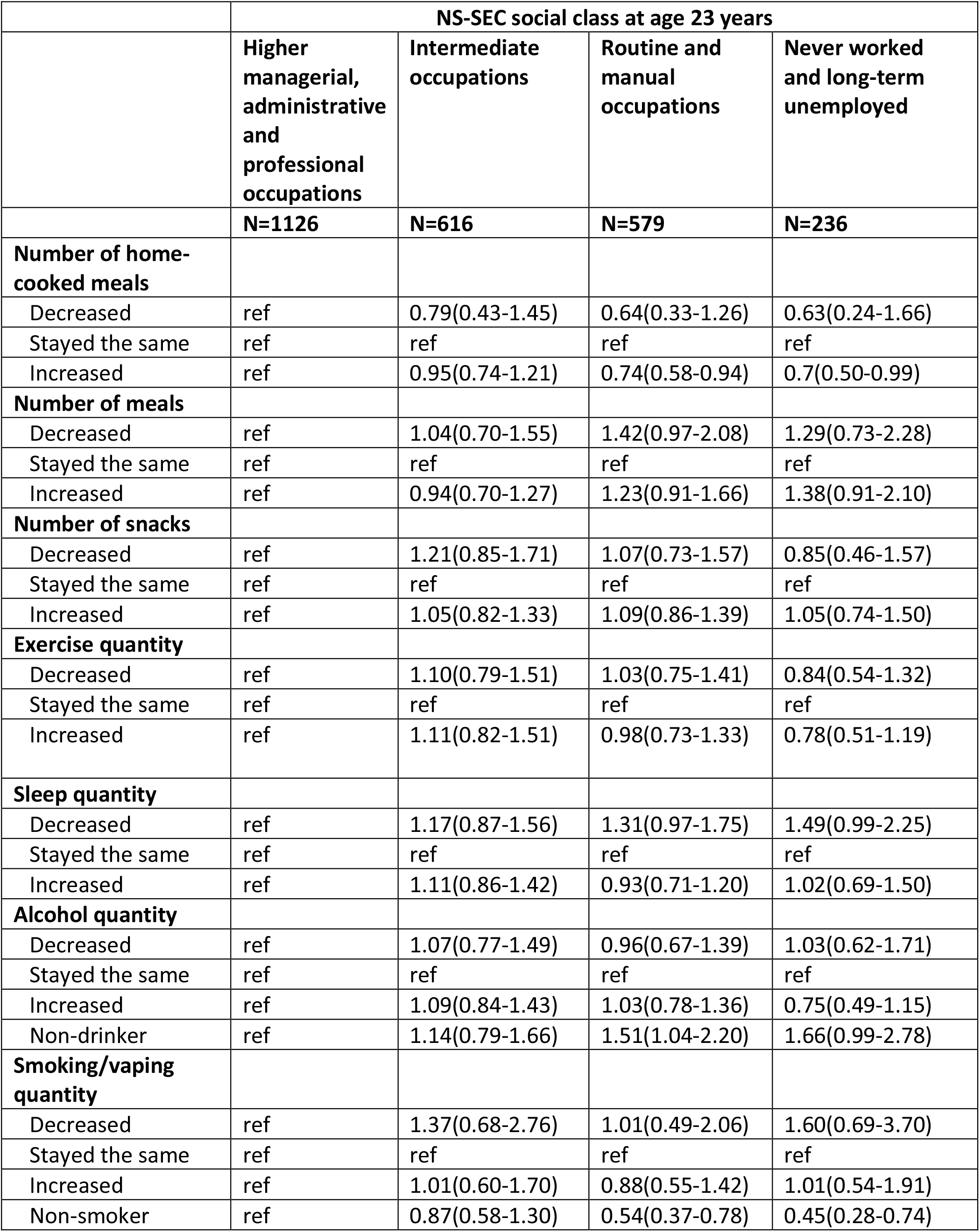
Association between social class and changes in health-related behaviour during the March-July 2020 lockdown (unadjusted). N=2557

|  | NS-SEC social class at age 23 years |  |  |  |
| --- | --- | --- | --- | --- |
|  | Higher managerial, administrative and professional occupations | Intermediate occupations | Routine and manual occupations | Never worked and long-term unemployed |
|  | N=1126 | N=616 | N=579 | N=236 |
| **Number of home-cooked meals** |  |  |  |  |
| Decreased | ref | 0.79(0.43-1.45) | 0.64(0.33-1.26) | 0.63(0.24-1.66) |
| Stayed the same | ref | ref | ref | ref |
| Increased | ref | 0.95(0.74-1.21) | 0.74(0.58-0.94) | 0.7(0.50-0.99) |
| **Number of meals** |  |  |  |  |
| Decreased | ref | 1.04(0.70-1.55) | 1.42(0.97-2.08) | 1.29(0.73-2.28) |
| Stayed the same | ref | ref | ref | ref |
| Increased | ref | 0.94(0.70-1.27) | 1.23(0.91-1.66) | 1.38(0.91-2.10) |
| **Number of snacks** |  |  |  |  |
| Decreased | ref | 1.21(0.85-1.71) | 1.07(0.73-1.57) | 0.85(0.46-1.57) |
| Stayed the same | ref | ref | ref | ref |
| Increased | ref | 1.05(0.82-1.33) | 1.09(0.86-1.39) | 1.05(0.74-1.50) |
| **Exercise quantity** |  |  |  |  |
| Decreased | ref | 1.10(0.79-1.51) | 1.03(0.75-1.41) | 0.84(0.54-1.32) |
| Stayed the same | ref | ref | ref | ref |
| Increased | ref | 1.11(0.82-1.51) | 0.98(0.73-1.33) | 0.78(0.51-1.19) |
| **Sleep quantity** |  |  |  |  |
| Decreased | ref | 1.17(0.87-1.56) | 1.31(0.97-1.75) | 1.49(0.99-2.25) |
| Stayed the same | ref | ref | ref | ref |
| Increased | ref | 1.11(0.86-1.42) | 0.93(0.71-1.20) | 1.02(0.69-1.50) |
| **Alcohol quantity** |  |  |  |  |
| Decreased | ref | 1.07(0.77-1.49) | 0.96(0.67-1.39) | 1.03(0.62-1.71) |
| Stayed the same | ref | ref | ref | ref |
| Increased | ref | 1.09(0.84-1.43) | 1.03(0.78-1.36) | 0.75(0.49-1.15) |
| Non-drinker | ref | 1.14(0.79-1.66) | 1.51(1.04-2.20) | 1.66(0.99-2.78) |
| **Smoking/vaping quantity** |  |  |  |  |
| Decreased | ref | 1.37(0.68-2.76) | 1.01(0.49-2.06) | 1.60(0.69-3.70) |
| Stayed the same | ref | ref | ref | ref |
| Increased | ref | 1.01(0.60-1.70) | 0.88(0.55-1.42) | 1.01(0.54-1.91) |
| Non-smoker | ref | 0.87(0.58-1.30) | 0.54(0.37-0.78) | 0.45(0.28-0.74) |

In the unadjusted model, exposure to 4+ ACEs was associated with reporting decreased sleep quantity during lockdown (odds ratio (OR) 1.53, 95% CI: 1.07-2.18) which attenuated slightly after adjustment for confounders (OR 1.39, 95% CI: 0.94-2.06) (Table 3; Supplementary Table 4). Experience of ACEs was related to change in both directions to number of meals per day. For example, people who had experienced 4+ ACEs were more than twice as likely to decrease (OR 2.34, 95% CI: 1.43-3.82) or increase (OR 2.18, 95% CI: 1.49-3.18) the number of meals compared with people reporting no ACEs. Experiencing 4+ ACEs was also associated with higher odds of increasing smoking and/or vaping (OR 1.85, 95% CI: 0.99-3.43). For other health behaviours, there was little evidence of associations with the ACE score.

**Table 3:**
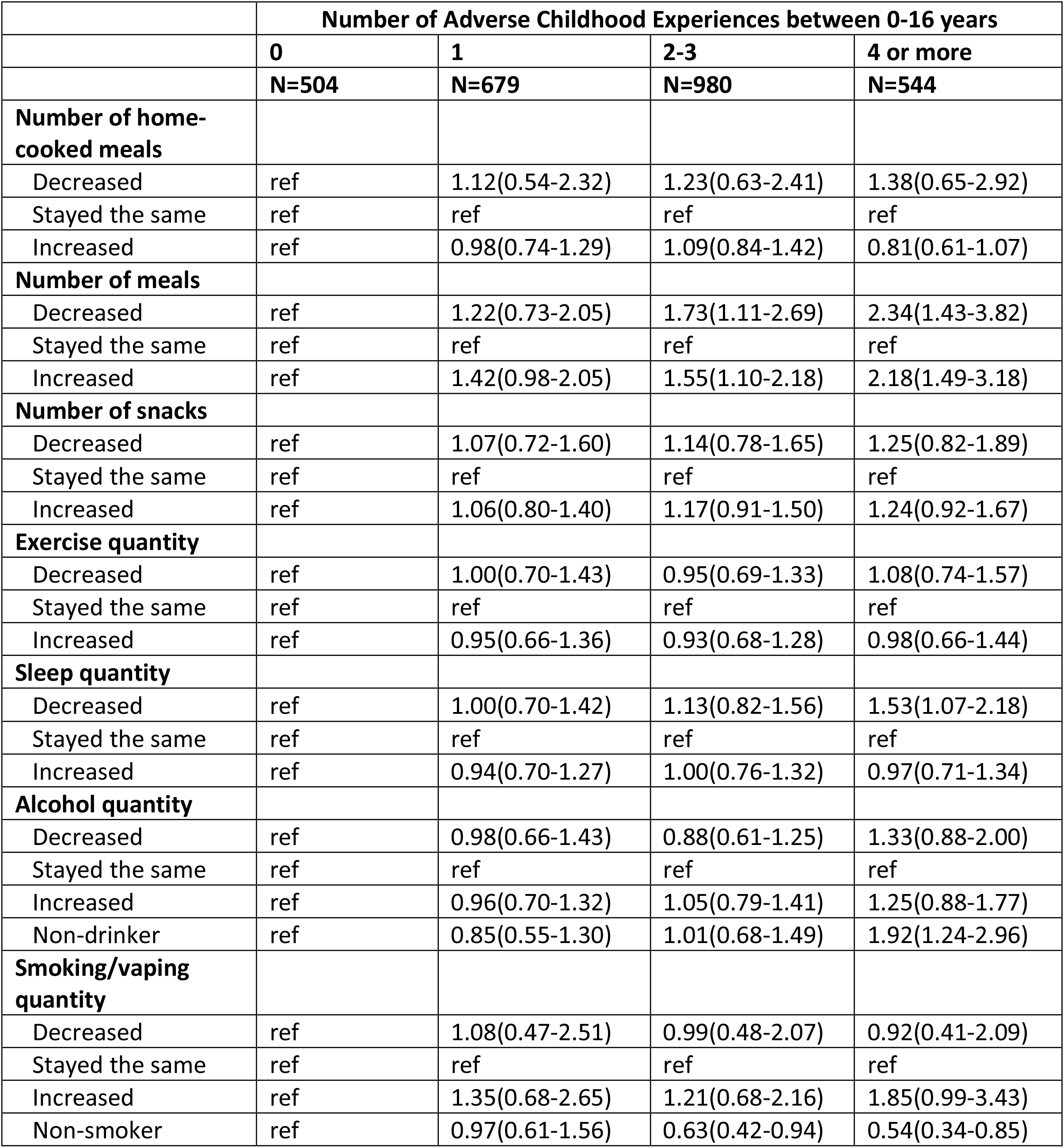
Association between Adverse Childhood Experiences score and changes in health-related behaviour during the March-July 2020 lockdown (unadjusted). N=2707

|  | **Number of Adverse Childhood Experiences between 0-16 years** |  |  |  |
| --- | --- | --- | --- | --- |
|  | **0** | **1** | **2-3** | **4 or more** |
|  | **N=504** | **N=679** | **N=980** | **N=544** |
| **Number of home-cooked meals** |  |  |  |  |
| Decreased | ref | 1.12(0.54-2.32) | 1.23(0.63-2.41) | 1.38(0.65-2.92) |
| Stayed the same | ref | ref | ref | ref |
| Increased | ref | 0.98(0.74-1.29) | 1.09(0.84-1.42) | 0.81(0.61-1.07) |
| **Number of meals** |  |  |  |  |
| Decreased | ref | 1.22(0.73-2.05) | 1.73(1.11-2.69) | 2.34(1.43-3.82) |
| Stayed the same | ref | ref | ref | ref |
| Increased | ref | 1.42(0.98-2.05) | 1.55(1.10-2.18) | 2.18(1.49-3.18) |
| **Number of snacks** |  |  |  |  |
| Decreased | ref | 1.07(0.72-1.60) | 1.14(0.78-1.65) | 1.25(0.82-1.89) |
| Stayed the same | ref | ref | ref | ref |
| Increased | ref | 1.06(0.80-1.40) | 1.17(0.91-1.50) | 1.24(0.92-1.67) |
| **Exercise quantity** |  |  |  |  |
| Decreased | ref | 1.00(0.70-1.43) | 0.95(0.69-1.33) | 1.08(0.74-1.57) |
| Stayed the same | ref | ref | ref | ref |
| Increased | ref | 0.95(0.66-1.36) | 0.93(0.68-1.28) | 0.98(0.66-1.44) |
| **Sleep quantity** |  |  |  |  |
| Decreased | ref | 1.00(0.70-1.42) | 1.13(0.82-1.56) | 1.53(1.07-2.18) |
| Stayed the same | ref | ref | ref | ref |
| Increased | ref | 0.94(0.70-1.27) | 1.00(0.76-1.32) | 0.97(0.71-1.34) |
| **Alcohol quantity** |  |  |  |  |
| Decreased | ref | 0.98(0.66-1.43) | 0.88(0.61-1.25) | 1.33(0.88-2.00) |
| Stayed the same | ref | ref | ref | ref |
| Increased | ref | 0.96(0.70-1.32) | 1.05(0.79-1.41) | 1.25(0.88-1.77) |
| Non-drinker | ref | 0.85(0.55-1.30) | 1.01(0.68-1.49) | 1.92(1.24-2.96) |
| **Smoking/vaping quantity** |  |  |  |  |
| Decreased | ref | 1.08(0.47-2.51) | 0.99(0.48-2.07) | 0.92(0.41-2.09) |
| Stayed the same | ref | ref | ref | ref |
| Increased | ref | 1.35(0.68-2.65) | 1.21(0.68-2.16) | 1.85(0.99-3.43) |
| Non-smoker | ref | 0.97(0.61-1.56) | 0.63(0.42-0.94) | 0.54(0.34-0.85) |

For individual ACEs, there was some evidence of associations with health behaviour changes (Supplementary Tables 5 & 6). Decreased sleep quantity during lockdown was associated with emotional abuse (OR 1.44, 95% CI: 1.09-1.90; unadjusted), physical abuse (OR 1.38, 95% CI: 1.07-1.78; unadjusted) and violence between parents (OR 1.38, 95% CI: 1.04-1.84; unadjusted). Participants with a history of violence between parents were twice as likely to increase smoking/vaping quantity during lockdown. For other individual ACEs, there was little evidence of associations with changes in health-related behaviours.

### Associations of SEP and ACEs with employment and financial situation during lockdown

People who were in NS-SEC group 3 or LTU at age 23 were more likely than those in NS-SEC group 1 at that age to experience the most adverse financial and employment outcomes (Table 4; Supplementary Table 7). For example, they were almost 3 times more likely to stop working during lockdown (OR 2.83, 95% CI: 1.45-5.50 for NS-SEC 3, OR 2.68, 95% CI: 1.63-4.42 for LTU). Both groups were also more than twice as likely to be on furlough or other leave, and about 50% more likely to have reduced employment hours.

**Table 4.** Association between social class and changes in financial situation during the March-July 2020 lockdown (unadjusted). N=2557

|  | **NS-SEC social class at age 23 years** |  |  |  |
| --- | --- | --- | --- | --- |
|  | **Higher managerial, administrative and professional occupations** | **Intermediate occupations** | **Routine and manual occupations** | **Never worked and long-term unemployed** |
|  | **N=1126** | **N=616** | **N=579** | **N=236** |
| **Change in employment** |  |  |  |  |
| Employed with the same or more hours | ref | ref | ref | ref |
| Reduced hours | ref | 0.89(0.59-1.36) | 1.51(1.01-2.24) | 1.65(0.95-2.87) |
| Furlough/paid or unpaid leave | ref | 1.41(1.04-1.92) | 2.53(1.86-3.43) | 2.20(1.40-3.47) |
| Stopped working during pandemic | ref | 1.00(0.59-1.67) | 2.54(1.58-4.08) | 2.57(1.35-4.87) |
| Not working pre-pandemic | ref | 1.28(0.77-2.13) | 2.34(1.45-3.79) | 5.40(3.20-9.12) |
| **Financial situation** |  |  |  |  |
| Worse off | ref | 0.97(0.73-1.29) | 1.39(1.05-1.84) | 1.36(0.89-2.06) |
| No change | ref | ref | ref | ref |
| Better off | ref | 0.95(0.74-1.22) | 0.73(0.56-0.97) | 0.73(0.49-1.10) |
| **Claimed benefits since pandemic** | ref | 0.96(0.66-1.40) | 1.39(0.97-1.98) | 1.96(1.22-3.16) |
| **Used rent/mortgage holidays since pandemic** | ref | 1.33(0.88-2.03) | 1.21(0.77-1.90) | 0.77(0.33-1.81) |

In the unadjusted model, an ACE score of 2-3 and 4+was associated with being furloughed or on paid or unpaid leave during lockdown (OR 1.45, 95% CI: 1.03-2.03 and OR 1.92, 95% CI: 1.35-2.74, respectively) (Table 5; Supplementary Table 8). Experiencing 4+ ACEs was also associated with a higher chance of stopping working during lockdown, but the confidence interval for this estimate was wide (OR=1.39, 95% CI 0.73-2.65). Experiencing 4+ ACEs was associated with feeling worse off financially during the pandemic, claiming new benefits, and using rent or mortgage holidays since the start of the pandemic. However estimates were imprecise, reflecting low prevalence of these outcomes.

**Table 5.** Association between Adverse Childhood Experiences score and changes in financial situation during the March-July 2020 lockdown (unadjusted). N=2707

|  | **Number of Adverse Childhood Experiences between 0-16 years** |  |  |  |
| --- | --- | --- | --- | --- |
|  | **0** | **1** | **2-3** | **4 or more** |
|  | **N=504** | **N=679** | **N=980** | **N=544** |
| **Change in employment** |  |  |  |  |
| Employed with the same or more hours | ref | ref | ref | ref |
| Reduced hours | ref | 0.82(0.53-1.29) | 1.03(0.69-1.55) | 0.99(0.62-1.57) |
| Furlough/paid or unpaid leave | ref | 1.01(0.70-1.46) | 1.45(1.03-2.03) | 1.92(1.35-2.74) |
| Stopped working during pandemic | ref | 1.03(0.58-1.80) | 1.31(0.78-2.21) | 1.39(0.73-2.65) |
| Not working pre-pandemic | ref | 1.11(0.65-1.91) | 1.31(0.78-2.19) | 2.11(1.22-3.66) |
| **Financial situation** |  |  |  |  |
| Worse off | ref | 0.97(0.68-1.38) | 1.26(0.92-1.71) | 1.42(1.00-2.02) |
| No change | ref | ref | ref | ref |
| Better off | ref | 0.86(0.65-1.14) | 0.87(0.66-1.14) | 0.83(0.60-1.15) |
| **Claimed benefits since pandemic** | ref | 1.36(0.90-2.06) | 1.14(0.77-1.68) | 1.36(0.87-2.12) |
| **Used rent/mortgage holidays since pandemic** | ref | 0.74(0.42-1.30) | 1.17(0.72-1.91) | 1.31(0.74-2.32) |

Some individual ACEs were associated with employment status change (Supplementary Table 9; Supplementary Table 10). For example, those with a history of physical abuse were more than twice as likely to have stopped working during lockdown than other participants (OR 2.15, 95% CI: 1.41-3.27). This was robust to adjustment for confounders (OR 2.32, 95% CI: 1.49-3.6). Participants with a history of parental substance abuse were almost twice as likely to have stopped working during lockdown compared to those without (OR 1.87, 95% CI: 1.05-3.33). Participants with a history of emotional neglect or parental separation were around 50% more likely to be furloughed or on other leave during lockdown than those without. A history of physical abuse or parental substance abuse was associated with being worse off financially during the pandemic. For other individual ACEs, there was little evidence of associations with changes in employment status or financial situation during lockdown.

## DISCUSSION

In a population-based longitudinal cohort study we identified changes in multiple health-related behaviours in both directions during lockdown, as well as changes to employment status and financial situation. The evidence of associations between SEP and health-related behaviour changes during lockdown was limited, but a higher overall ACE score and various individual ACEs were found to be associated with some behaviours e.g., decreased sleep and increased smoking. There was, however, clear evidence that adverse employment and financial changes were more likely to be experienced by people with low SEP or a history of ACE exposure. In this cohort of young adults, people in routine or manual occupations or long-term unemployment four years prior to the pandemic were more likely to experience adverse changes to their employment status during lockdown compared to participants in higher managerial, administrative, or professional occupations four years previous. Those with a higher ACE score were more likely to be put on furlough or other leave or stop working entirely during lockdown. In addition, we found some evidence that the group with the greatest ACE exposure were more likely to experience adverse financial outcomes during lockdown.

There is a growing body of evidence of the indirect effects of lockdown on health-related behaviours. A study analysing five British cohorts of different ages investigated whether behaviour change during lockdown differed by SEP and, like the present study, found that adverse effects on sleep during lockdown were more frequent amongst socioeconomically disadvantaged groups.[7] The same study, which had a large combined sample size, found that lower SEP was associated with lower exercise quantity during lockdown, something that wasn’t detected in our sample. Analysis of a large UK cohort found that overall smoking declined during lockdown and that there was no interaction of smoking behaviour with education level.[11] There is evidence of socioeconomic disparities in drinking behaviours during lockdown,[13,14] which we did not detect in our sample.

In keeping with our findings, adverse effects on employment seem to be more common for younger people and those in lower paid occupations.^6,18,19^ One study exploring adversities during the first 3 weeks of lockdown by SEP found that people of low SEP were more likely to lose work, experience a cut in household income and be unable to pay bills during lockdown than people of higher SEP.^19^ Similarly, a Welsh study found that unemployment and furlough during lockdown disproportionately affected those experiencing financial difficulties and living in deprived communities.^20^ To our knowledge, ours is the first study to assess ACEs as a risk factor for adverse employment and financial changes during lockdown.

This study has limitations. The sample size and thus statistical power was restricted as the questionnaire was online only - invites were only sent to those participants for whom the study had a valid email address, impacting participants who prefer to complete questionnaires on paper. Self-reported measures were used, so associations may have been biased by measurement error and reporting biases. ALSPAC participants are more socioeconomically advantaged and less ethnically diverse than the national average, and mothers of participants included in this study had higher educational attainment compared to the rest of the ALSPAC cohort. Results therefore may not be generalisable to the UK population. Although ALSPAC participants are in the age group most likely to be affected by lockdown in terms of employment^18^, their similar ages meant we could not study the effect of age on the outcomes. Finally, our SEP measure was solely based on participants’ occupations, but individual’s income, education, or residential area may also determine negative impacts of lockdown.

### Conclusion

The results of this study support findings from routine data that the financial and employment situation of adults with lower occupational social class are more likely to have been adversely affected by the COVID-19 non-pharmaceutical interventions.[35] However, importantly, our findings also highlight the economic adversity experienced by adults who have been exposed to ACEs. Despite the long-reaching consequences of ACEs persisting into adulthood, there are few interventions and services targeted at this group (with most services for adults targeting adult risk factors rather than previous childhood risk factors), and they are largely hidden from view in routine data. Our findings highlight a need for continued support of people who experience ACEs into adulthood, and demonstrate that this need may have increased during the COVID-19 pandemic.

Our results also suggest that exposure to multiple ACEs was associated with decreased sleep quantity and increased smoking/vaping, but we found little evidence that SEP or ACEs influenced changes in other health-related behaviours during the pandemic in this study of young adults, providing some reassurance that, at least in the short-term, the non-pharmaceutical interventions used against COVID-19 have not led to increased inequalities in health-related behaviours in this age group.

## Data Availability

ALSPAC data are available by application to the study executive committee, subject to submission and approval of a project proposal and payment of data access fees. For details and inquiries, please see http://www.bristol.ac.uk/alspac/researchers/access/ or. Full details of all ALSPAC data are available in the ALSPAC data dictionary, which can be found at http://www.bristol.ac.uk/alspac/external/documents/ALSPAC_Data_Dictionary.zip.

http://www.bristol.ac.uk/alspac/external/documents/ALSPAC_Data_Dictionary.zip

## ACKNOWLEDGEMENTS

We are extremely grateful to all the families who took part in this study, the midwives for their help in recruiting them, and the whole ALSPAC team, which includes interviewers, computer and laboratory technicians, clerical workers, research scientists, volunteers, managers, receptionists and nurses.

## CONTRIBUTORS

LDH and A Hughes conceived the idea for the study. MLS and A Herbert conducted the analysis. MLS drafted the manuscript. MLS, LDH, A Hughes, A Herbert and KN contributed to the study design, interpretation of the findings and critically revised the manuscript. All authors approved the final version of the paper.

---

**Supplementary Figure 1:**
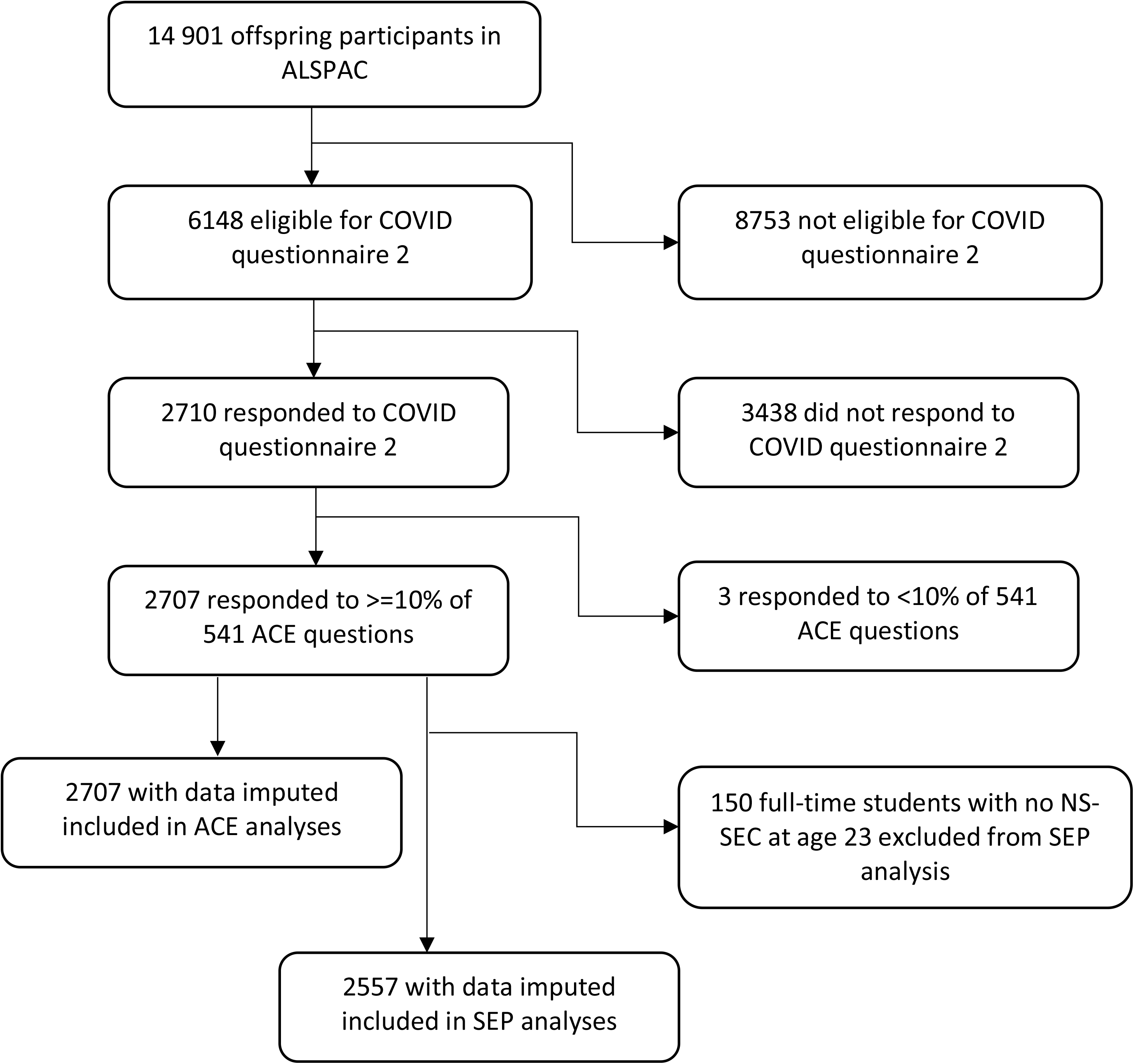
Flow chart of inclusion and exclusion of participants

**Supplementary Table 1.**
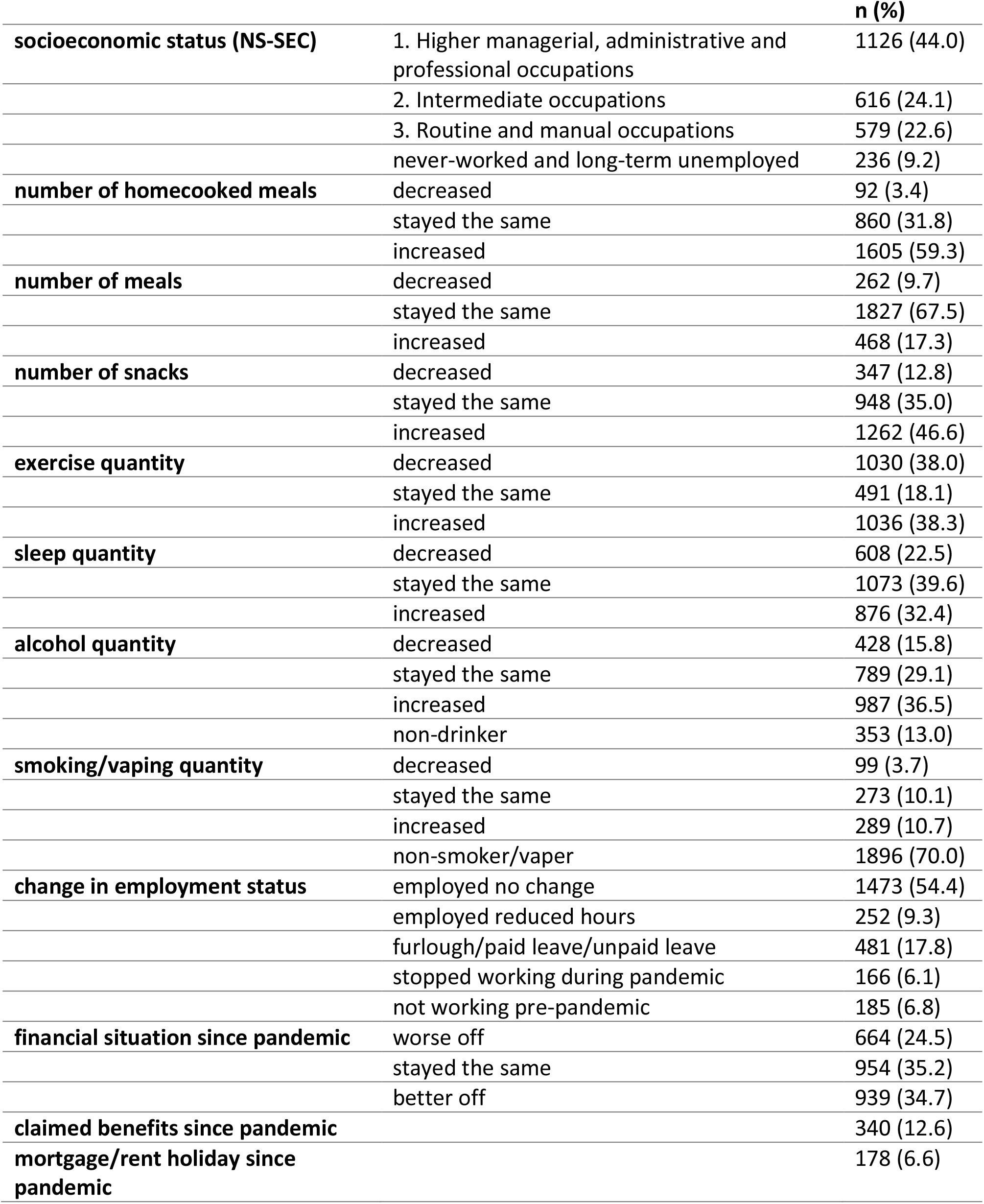
Participant characteristics. Characteristics of the participants included in SEP analyses using data from multi-variate multiple imputation. N=2557.

**Supplementary Table 2.** Maternal education for excluded participants and participants included in analyses.

|  | **Participants (%)** |  |
| --- | --- | --- |
|  | **Excluded** | **Included** |
|  | N=12911 | N=2710 |
| **Mother's highest education qualification** |  |  |
| CSE | 13.6 | 8.2 |
| Vocational | 7.0 | 6.2 |
| O level | 24.3 | 30.3 |
| A level | 14.9 | 25.1 |
| Degree | 7.6 | 20.1 |
| Not available | 32.6 | 10.1 |

**Supplementary Table 3:** Association between social class and changes in health-related behaviour during the March-July 2020 lockdown (adjusted for ethnicity, age at time of questionnaire, home ownership, maternal and partner education, parity, maternal age and maternal marital status). N=2557

|  | NS-SEC social class at age 23 years |  |  |  |
| --- | --- | --- | --- | --- |
|  | Higher managerial, administrative and professional occupations | Intermediate occupations | Routine and manual occupations | Never worked and long-term unemployed |
|  | N=1126 | N=616 | N=579 | N=236 |
| **Number of home-cooked meals** |  |  |  |  |
| Decreased | ref | 0.79(0.42-1.47) | 0.63(0.31-1.28) | 0.58(0.21-1.57) |
| Stayed the same | ref | ref | ref | ref |
| Increased | ref | 0.97(0.76-1.25) | 0.77(0.59-1.00) | 0.72(0.51-1.04) |
| **Number of meals** |  |  |  |  |
| Decreased | ref | 1.03(0.69-1.54) | 1.41(0.95-2.09) | 1.22(0.67-2.23) |
| Stayed the same | ref | ref | ref | ref |
| Increased | ref | 0.91(0.67-1.24) | 1.13(0.83-1.55) | 1.26(0.81-1.97) |
| **Number of snacks** |  |  |  |  |
| Decreased | ref | 1.23(0.86-1.76) | 1.09(0.73-1.62) | 0.85(0.45-1.59) |
| Stayed the same | ref | ref | ref | ref |
| Increased | ref | 1.02(0.80-1.31) | 1.02(0.80-1.31) | 0.99(0.69-1.43) |
| **Exercise quantity** |  |  |  |  |
| Decreased | ref | 1.08(0.77-1.50) | 1.02(0.73-1.42) | 0.83(0.52-1.33) |
| Stayed the same | ref | ref | ref | ref |
| Increased | ref | 1.11(0.81-1.52) | 0.99(0.72-1.37) | 0.78(0.50-1.23) |
| **Sleep quantity** |  |  |  |  |
| Decreased | ref | 1.16(0.86-1.55) | 1.26(0.93-1.70) | 1.41(0.92-2.16) |
| Stayed the same | ref | ref | ref | ref |
| Increased | ref | 1.12(0.87-1.44) | 0.99(0.75-1.30) | 1.11(0.73-1.67) |
| **Alcohol quantity** |  |  |  |  |
| Decreased | ref | 1.09(0.78-1.52) | 1.00(0.68-1.47) | 1.02(0.61-1.72) |
| Stayed the same | ref | ref | ref | ref |
| Increased | ref | 1.08(0.82-1.42) | 1.08(0.82-1.42) | 0.74(0.48-1.16) |
| Non-drinker | ref | 1.08(0.74-1.59) | 1.36(0.92-2.02) | 1.42(0.83-2.44) |
| **Smoking/vaping quantity** |  |  |  |  |
| Decreased | ref | 1.39(0.68-2.84) | 1.11(0.53-2.35) | 1.68(0.7-4.04) |
| Stayed the same | ref | ref | ref | ref |
| Increased | ref | 0.97(0.57-1.65) | 0.86(0.53-1.41) | 0.93(0.47-1.82) |
| Non-smoker | ref | 0.89(0.59-1.34) | 0.58(0.40-0.86) | 0.48(0.29-0.79) |

**Supplementary Table 4:** Association between Adverse Childhood Experiences score and changes in health-related behaviour during the March-July 2020 lockdown (adjusted for ethnicity, age at time of questionnaire, home ownership, maternal and partner education, parity, maternal age and maternal marital status). N=2707

|  | Number of Adverse Childhood Experiences between 0-16 years |  |  |  |
| --- | --- | --- | --- | --- |
|  | 0 | 1 | 2-3 | 4 or more |
|  | N=504 | N=679 | N=980 | N=544 |
| **Number of home-cooked meals** |  |  |  |  |
| Decreased | ref | 1.15(0.55-2.41) | 1.25(0.63-2.50) | 1.44(0.64-3.24) |
| Stayed the same | ref | ref | ref | ref |
| Increased | ref | 0.99(0.74-1.31) | 1.16(0.89-1.51) | 0.87(0.64-1.19) |
| **Number of meals** |  |  |  |  |
| Decreased | ref | 1.21(0.72-2.04) | 1.59(1.01-2.51) | 1.90(1.12-3.23) |
| Stayed the same | ref | ref | ref | ref |
| Increased | ref | 1.34(0.92-1.95) | 1.35(0.95-1.91) | 1.61(1.07-2.41) |
| **Number of snacks** |  |  |  |  |
| Decreased | ref | 1.10(0.73-1.64) | 1.17(0.80-1.72) | 1.21(0.76-1.91) |
| Stayed the same | ref | ref | ref | ref |
| Increased | ref | 1.02(0.77-1.36) | 1.08(0.84-1.40) | 1.06(0.76-1.47) |
| **Exercise quantity** |  |  |  |  |
| Decreased | ref | 1.02(0.71-1.48) | 0.99(0.70-1.40) | 1.08(0.71-1.65) |
| Stayed the same | ref | ref | ref | ref |
| Increased | ref | 0.96(0.67-1.39) | 0.98(0.71-1.36) | 1.06(0.70-1.63) |
| **Sleep quantity** |  |  |  |  |
| Decreased | ref | 1.00(0.70-1.43) | 1.08(0.77-1.50) | 1.39(0.94-2.06) |
| Stayed the same | ref | ref | ref | ref |
| Increased | ref | 0.97(0.72-1.32) | 1.06(0.80-1.41) | 1.06(0.75-1.52) |
| **Alcohol quantity** |  |  |  |  |
| Decreased | ref | 0.98(0.66-1.45) | 0.90(0.62-1.30) | 1.39(0.89-2.16) |
| Stayed the same | ref | ref | ref | ref |
| Increased | ref | 0.98(0.71-1.34) | 1.08(0.80-1.46) | 1.34(0.92-1.96) |
| Non-drinker | ref | 0.81(0.52-1.24) | 0.88(0.59-1.32) | 1.55(0.95-2.53) |
| **Smoking/vaping quantity** |  |  |  |  |
| Decreased | ref | 1.05(0.45-2.48) | 0.99(0.47-2.10) | 1.00(0.41-2.45) |
| Stayed the same | ref | ref | ref | ref |
| Increased | ref | 1.35(0.68-2.68) | 1.18(0.65-2.14) | 1.82(0.94-3.51) |
| Non-smoker | ref | 0.99(0.61-1.60) | 0.67(0.44-1.00) | 0.63(0.38-1.04) |

**Supplementary Table 5:**
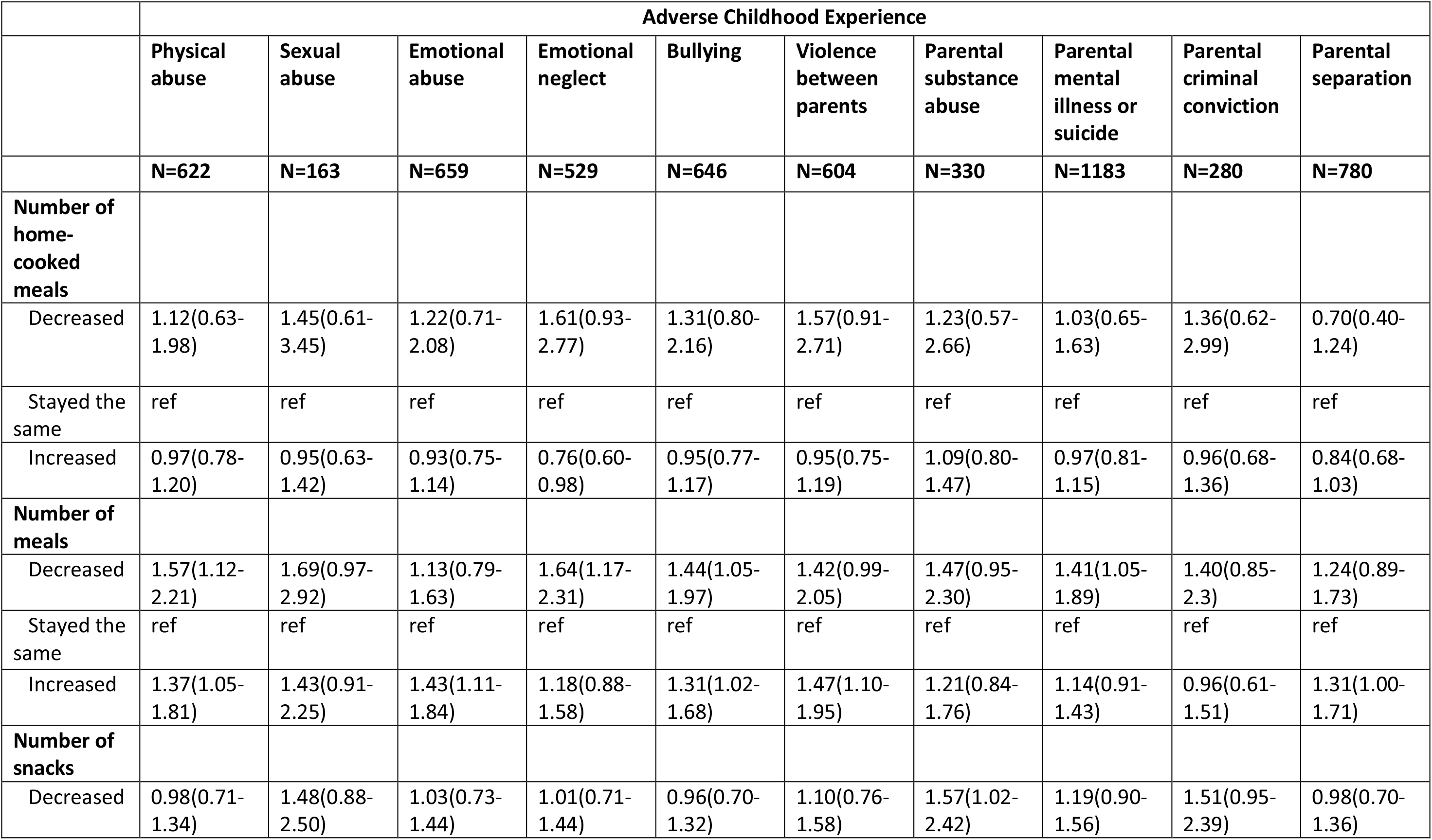
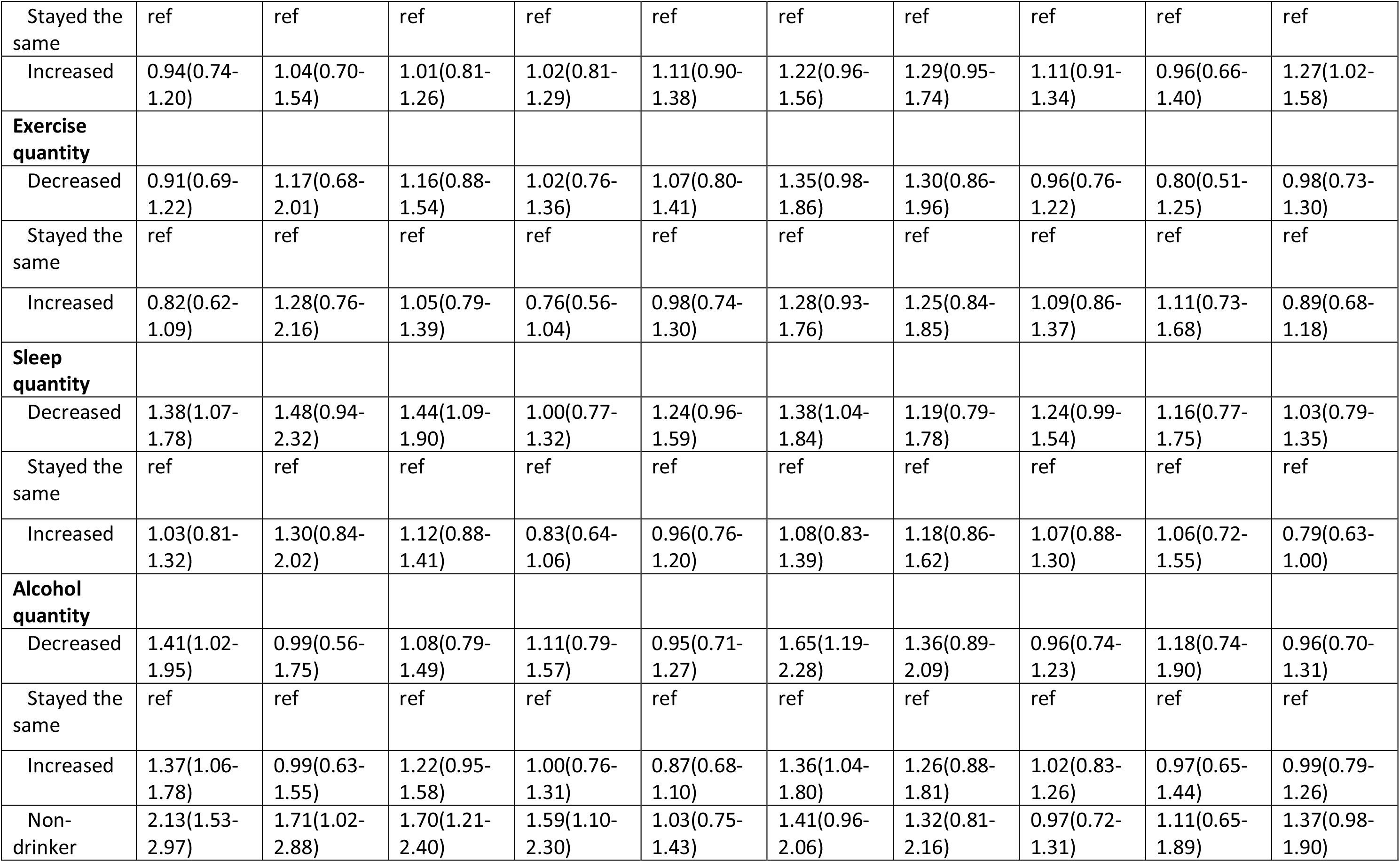
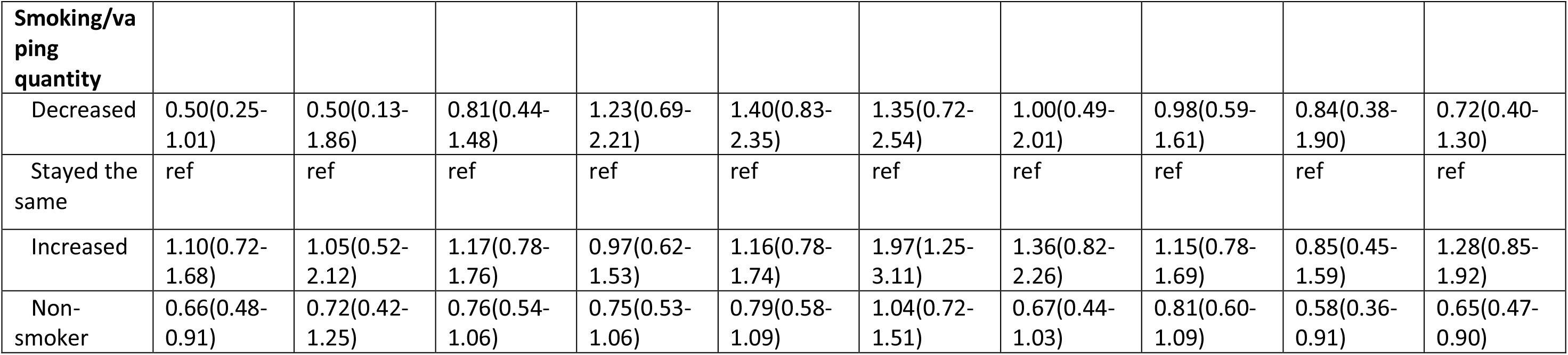
Association between individual Adverse Childhood Experiences and changes in health-related behaviour during the March-July 2020 lockdown (unadjusted). N=2707

**Supplementary Table 6:**
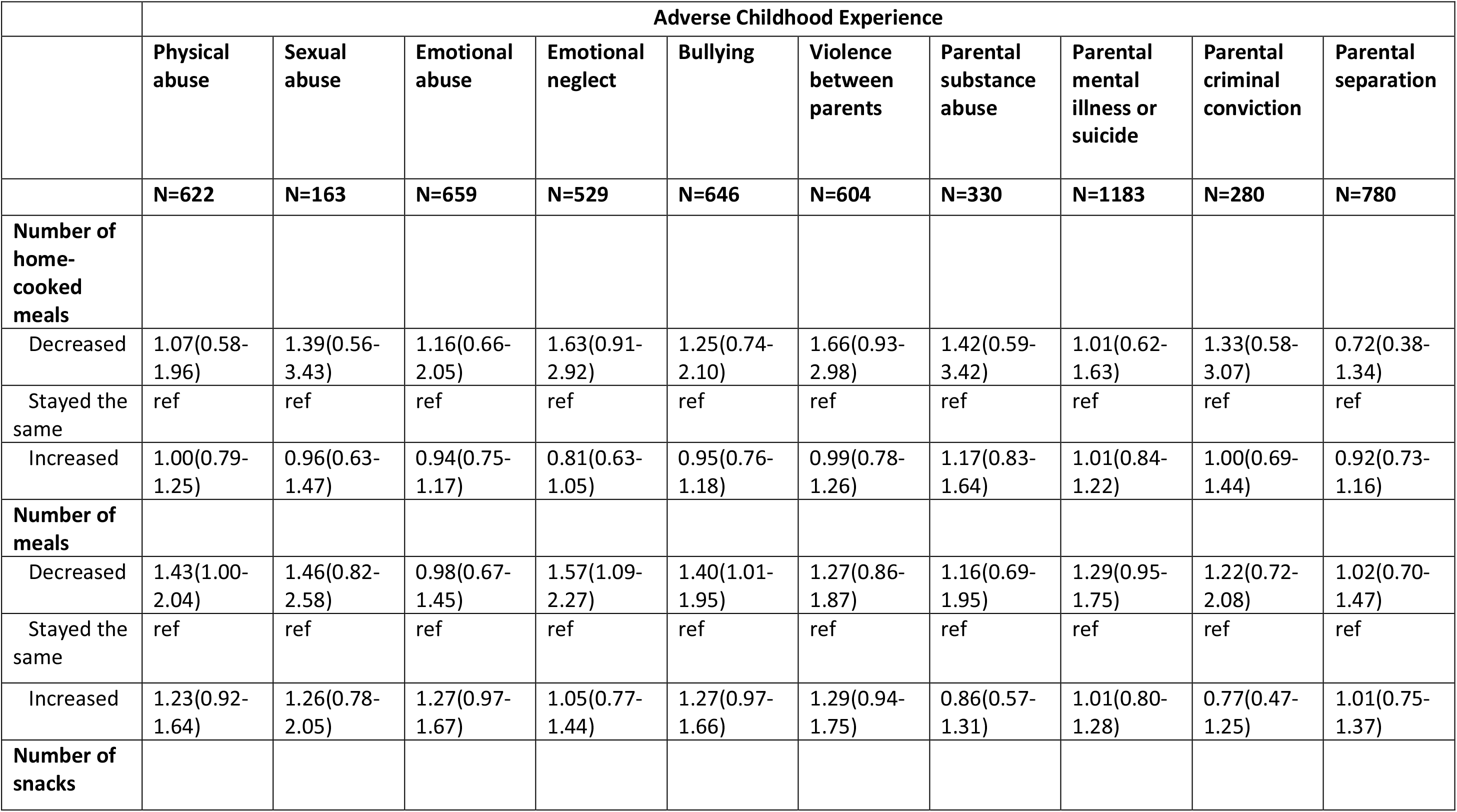
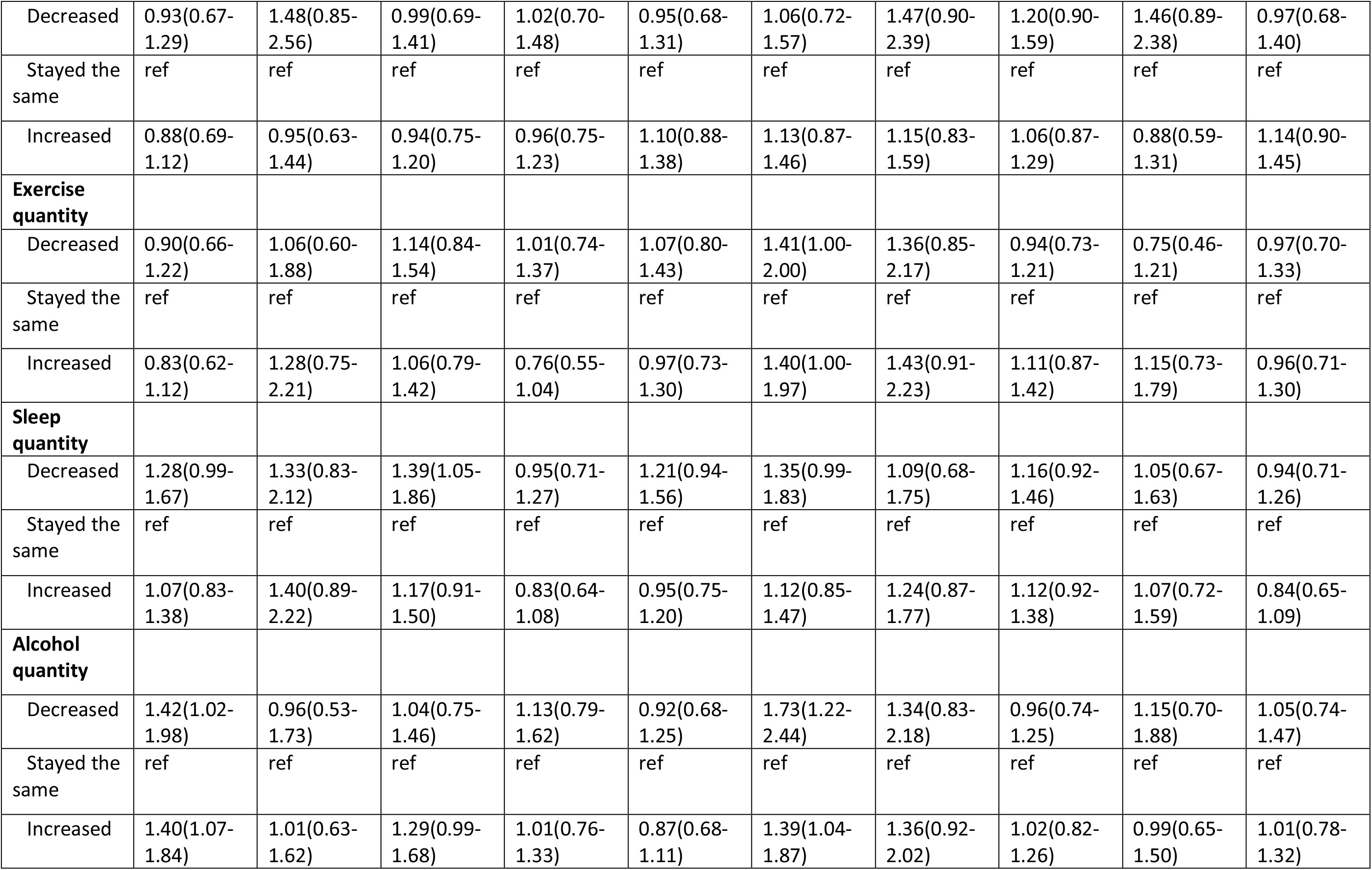
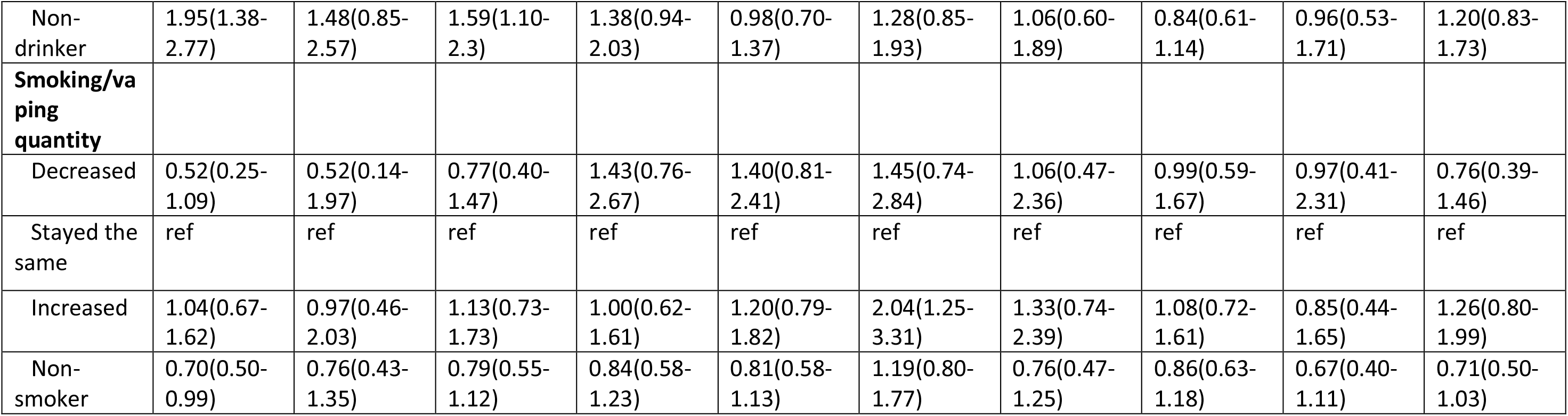
Association between individual Adverse Childhood Experiences and changes in health-related behaviour during the March-July 2020 lockdown (adjusted for ethnicity, age at time of questionnaire, home ownership, maternal and partner education, parity, maternal age and maternal marital status). N=2707

**Supplementary Table 7.** Association between social class and changes in financial situation during the March-July 2020 lockdown (adjusted for ethnicity, age at time of questionnaire, home ownership, maternal and partner education, parity, maternal age and maternal marital status). N=2557

|  | NS-SEC social class at age 23 years |  |  |  |
| --- | --- | --- | --- | --- |
|  | Higher managerial, administrative and professional occupations | Intermediate occupations | Routine and manual occupations | Never worked and long-term unemployed |
|  | N=1126 | N=616 | N=579 | N=236 |
| **Change in employment** |  |  |  |  |
| Employed with the same or more hours | ref | ref | ref | ref |
| Reduced hours | ref | 0.87(0.57-1.34) | 1.45(0.96-2.18) | 1.54(0.87-2.75) |
| Furlough/paid or unpaid leave | ref | 1.39(1.02-1.91) | 2.38(1.73-3.26) | 2.10(1.30-3.38) |
| Stopped working during pandemic | ref | 0.98(0.58-1.67) | 2.68(1.63-4.42) | 2.83(1.45-5.50) |
| Not working pre-pandemic | ref | 1.21(0.72-2.05) | 2.11(1.27-3.51) | 5.38(3.05-9.49) |
| **Financial situation** |  |  |  |  |
| Worse off | ref | 0.98(0.73-1.31) | 1.45(1.08-1.96) | 1.45(0.95-2.23) |
| No change | ref | ref | ref | ref |
| Better off | ref | 0.99(0.76-1.29) | 0.81(0.61-1.08) | 0.81(0.53-1.23) |
| **Claimed benefits since pandemic** | ref | 0.95(0.65-1.41) | 1.45(0.99-2.10) | 2.06(1.24-3.43) |
| **Used rent/mortgage holidays since pandemic** | ref | 1.28(0.83-1.97) | 1.15(0.72-1.85) | 0.73(0.30-1.79) |

**Supplementary Table 8.** Association between Adverse Childhood Experiences score and changes in financial situation during the March-July 2020 lockdown (adjusted for ethnicity, age at time of questionnaire, home ownership, maternal and partner education, parity, maternal age and maternal marital status). N=2707

|  | Number of Adverse Childhood Experiences between 0-16 years |  |  |  |
| --- | --- | --- | --- | --- |
|  | 0 | 1 | 2-3 | 4 or more |
|  | N=504 | N=679 | N=980 | N=544 |
| **Change in employment** |  |  |  |  |
| Employed with the same or more hours | ref | ref | ref | ref |
| Reduced hours | ref | 0.80(0.50-1.27) | 0.95(0.62-1.45) | 0.83(0.49-1.38) |
| Furlough/paid or unpaid leave | ref | 0.96(0.66-1.41) | 1.28(0.90-1.81) | 1.51(1.02-2.24) |
| Stopped working during pandemic | ref | 1.00(0.57-1.77) | 1.28(0.75-2.18) | 1.37(0.68-2.78) |
| Not working pre-pandemic | ref | 1.06(0.61-1.84) | 1.16(0.68-1.98) | 1.66(0.91-3.01) |
| **Financial situation** |  |  |  |  |
| Worse off | ref | 0.98(0.69-1.39) | 1.26(0.92-1.72) | 1.45(0.98-2.14) |
| No change | ref | ref | ref | ref |
| Better off | ref | 0.89(0.67-1.19) | 0.95(0.71-1.26) | 0.93(0.66-1.31) |
| **Claimed benefits since pandemic** | ref | 1.37(0.90-2.08) | 1.13(0.75-1.69) | 1.32(0.81-2.15) |
| **Used rent/mortgage holidays since pandemic** | ref | 0.72(0.41-1.28) | 1.10(0.67-1.82) | 1.19(0.64-2.22) |

**Supplementary Table 9.** Association between individual Adverse Childhood Experiences and changes in financial situation during the March-July 2020 lockdown (unadjusted). N=2707

|  | Adverse Childhood Experience |  |  |  |  |  |  |  |  |  |
| --- | --- | --- | --- | --- | --- | --- | --- | --- | --- | --- |
|  | Physical abuse | Sexual abuse | Emotional abuse | Emotional neglect | Bullying | Violence between parents | Parental substance abuse | Parental mental illness or suicide | Parental criminal conviction | Parental separation |
|  | N=622 | N=163 | N=659 | N=529 | N=646 | N=604 | N=330 | N=1183 | N=280 | N=780 |
| Change in employment |  |  |  |  |  |  |  |  |  |  |
| Employed with the same or more hours | ref | ref | ref | ref | ref | ref | ref | ref | ref | ref |
| Reduced hours | 1.02(0.70-1.48) | 1.27(0.69-2.32) | 1.03(0.71-1.50) | 1.41(0.98-2.03) | 1.15(0.79-1.67) | 0.74(0.47-1.15) | 1.52(0.95-2.42) | 0.87(0.64-1.18) | 0.77(0.41-1.46) | 1.08(0.75-1.54) |
| Furlough/ paid or unpaid leave | 1.30(0.99-1.72) | 1.61(1.02-2.55) | 1.29(0.99-1.69) | 1.44(1.08-1.92) | 1.29(0.99-1.68) | 1.41(1.06-1.89) | 1.39(0.95-2.04) | 1.09(0.87-1.37) | 1.20(0.78-1.83) | 1.48(1.16-1.90) |
| Stopped working during pandemic | 2.15(1.41-3.27) | 0.97(0.41-2.32) | 1.13(0.70-1.81) | 0.95(0.55-1.63) | 1.19(0.79-1.81) | 0.84(0.47-1.51) | 1.87(1.05-3.33) | 1.02(0.70-1.46) | 1.01(0.50-2.02) | 0.84(0.52-1.37) |
| Not working pre-pandemic | 1.43(0.96-2.13) | 1.23(0.61-2.48) | 1.19(0.78-1.82) | 1.75(1.16-2.66) | 1.31(0.90-1.92) | 1.27(0.81-1.99) | 1.15(0.62-2.13) | 1.28(0.92-1.80) | 0.85(0.42-1.72) | 1.53(1.05-2.23) |
| **Financial situation** |  |  |  |  |  |  |  |  |  |  |
| Worse off | 1.35(1.03-1.77) | 1.41(0.88-2.25) | 1.05(0.79-1.38) | 0.97(0.74-1.28) | 1.12(0.86-1.46) | 1.04(0.78-1.37) | 1.45(1.02-2.08) | 1.20(0.96-1.51) | 1.32(0.88-1.99) | 1.18(0.91-1.52) |
| No change | ref | ref | ref | ref | ref | ref | ref | ref | ref | ref |
| Better off | 0.91(0.71-1.16) | 1.20(0.77-1.85) | 1.08(0.84-1.38) | 0.81(0.62-1.05) | 0.93(0.74-1.18) | 0.86(0.67-1.10) | 1.14(0.81-1.62) | 1.01(0.82-1.24) | 1.28(0.86-1.92) | 0.89(0.70-1.13) |
| **Claimed benefits since pandemic** | 1.15(0.85-1.56) | 1.21(0.70-2.08) | 1.09(0.79-1.50) | 0.95(0.67-1.35) | 1.11(0.80-1.52) | 0.93(0.65-1.35) | 1.35(0.89-2.05) | 0.98(0.75-1.28) | 1.10(0.67-1.81) | 0.99(0.73-1.33) |
| **Used rent/mortgage holidays since pandemic** | 1.16(0.77-1.75) | 1.13(0.52-2.45) | 0.90(0.56-1.44) | 1.02(0.62-1.70) | 1.23(0.83-1.81) | 1.47(0.96-2.24) | 1.16(0.65-2.08) | 1.23(0.86-1.76) | 1.23(0.66-2.29) | 1.43(0.96-2.13) |

**Supplementary Table 10.**
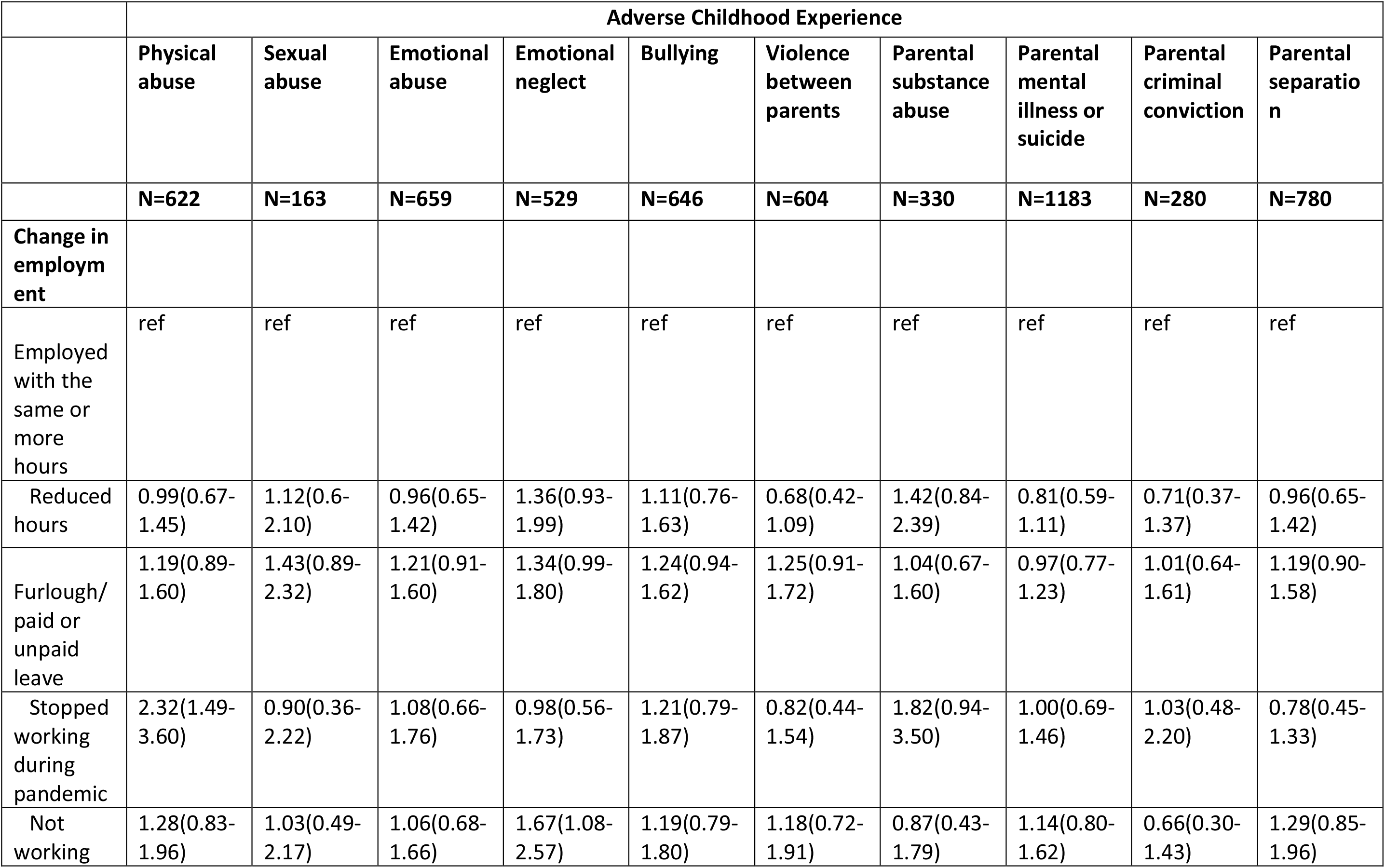
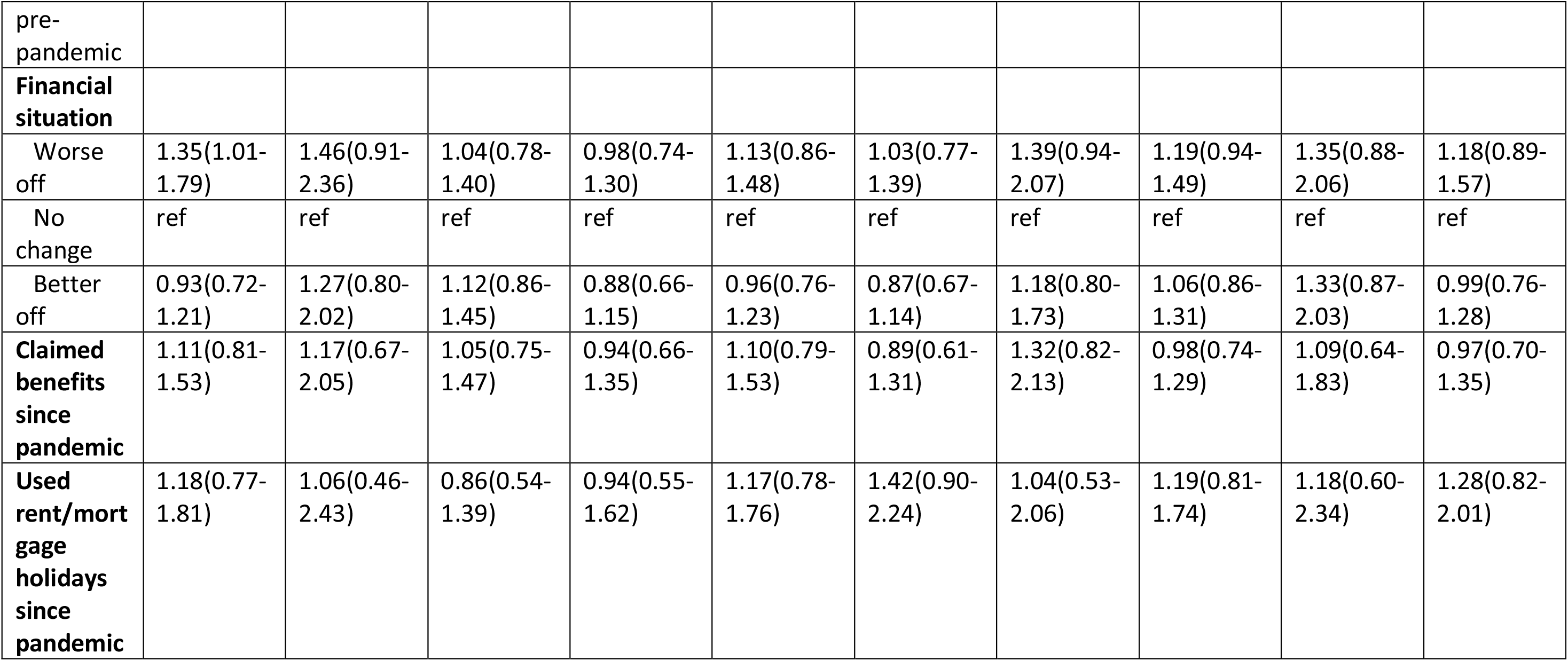
Association between individual Adverse Childhood Experiences and changes in financial situation during the March-July 2020 lockdown (adjusted for ethnicity, age at time of questionnaire, home ownership, maternal and partner education, parity, maternal age and maternal marital status). N=2707

