## Supplementary File for "Associations of socioeconomic position and adverse childhood experiences with health-related behaviour changes and changes to employment during the first COVID-19 lockdown in the UK"

**Additional methodological details**

**Dichotomous ACE indicators**

Multiple questions fed into each ACE indicator. If an ACE was reported in one or more question, it was assumed to have been experienced, even if there were inconsistencies in responses. ACE indicators were mostly derived from data collected prospectively, with the inclusion of some retrospective self-reported measures, particularly for sexual abuse where the reported prevalence in prospective data is very low.

**Auxiliary variables for multivariate multiple imputation**

Of 110 possible auxiliary variables, the following 24 had at least 50 observations within each level of that variable for both males and females, and so were used in the multiple imputation models (terms in brackets refer to the original ALSPAC variable labels):

- Birthweight (kz030_org)
- Gestation (kz029_org)
- Pre-pregnancy weight and BMI (dw002_org, dw042_org)
- Home ownership status (a006_org)
- Mother’s age at delivery (mz028b_org)
- Parity (b032_org)
- Mother’s marital status (a525_org)
- Mother’s and partner’s highest educational qualifications (c645a_org, c666a_org, pb325a_org, pb342a_org)
- Indicators of mother’s and partner’s mental health conditions (b370_org, c600_org, pb260_org, t3255_org, t5360_org, t5404)
- In last year, father became homeless (fa3322_org)
- Mother or partner smoked, used cannabis/hard drugs or suffered alcoholism (t5412_org, t5510_org, fa5411_org, fa5510_org)
- Young person’s age when partners have used physical force such as pushing, slapping, hitting or holding them down (ypa5005_dup)
